# Characterization of prolonged COVID-19 symptoms and patient comorbidities in an outpatient telemedicine cohort

**DOI:** 10.1101/2020.07.05.20146886

**Authors:** Michele Cellai, James B. O’Keefe

## Abstract

We identified patients with coronavirus disease 2019 (COVID-19) in a telemedicine clinic who requested ongoing follow-up 6 weeks after symptom onset. Patients with prolonged symptoms often have not returned to work or usual activity. Respiratory symptoms are most common, and underlying asthma and lung disease occur frequently.

## Introduction

While the early clinical course of ambulatory patients with coronavirus disease 2019 (COVID-19) is rapidly being described [1-4], data on symptoms persisting 5 weeks or longer are limited. Data illustrating delayed recovery are available from the severe acute respiratory virus disease (SARS) outbreak in 2003 [4] as well as community-acquired pneumonia [5], but these reports are limited to post-hospitalization cohorts. Anecdotally, a subset of patients with COVID-19 reported delayed return to normal activity, even among patients with mild illness managed at home.

The Emory Clinic Virtual Outpatient Management Clinic (VOMC) is a telemedicine program for patients with COVID-19 diagnosed by nasopharyngeal polymerase chain reaction (PCR) during their time in isolation at home. The VOMC follows patients with confirmed COVID-19 with regular telephone calls (for 7-21 days depending on patient symptom severity, comorbidities, and age). The calls may be continued at the request of the patient and/or provider (registered nurse or advanced practice provider). We seek to describe the persistent symptoms experienced by patients with mild COVID-19 at 5 or more weeks of illness by reviewing records of patients who continued to request follow-up care for greater than 6 weeks beyond symptom onset. We hypothesize that specific comorbidities or age may be associated with prolonged symptom duration in comparison to the overall VOMC population.

## Methods

We conducted a search of the patients enrolled in the VOMC between March 24 and May 26, 2020 who met inclusion criteria: (1) enrollment in VOMC within 6 days of a positive COVID-19 PCR, (2) specific symptom onset date recorded at the time of VOMC intake, and (3) final VOMC follow-up call greater than 6 weeks after symptom onset date. Exclusion criteria included: (1) hospitalization prior to entering VOMC, (2) VOMC care duration less than 3 weeks. Charts were reviewed through June 8, at which time all eligible patients based on inclusion criteria had been discharged from VOMC.

Chart review included (1) verification of patient demographics and comorbidities, (2) verification of symptom onset dates (from screening clinic note and VOMC intake note), (3) review of follow-up notes during the week 5-6 time range, (4) review of return to work/disability letters, and (5) review of final notes for discharge care plan. For patients with significant symptoms requiring medical evaluation (in-person acute visit or telemedicine visit with primary care physician or specialist) at least 21 days into symptom course, we reviewed evaluation notes, diagnostics, and final diagnosis (recorded as either alternate diagnosis unrelated to COVID-19 or suspect contributing diagnosis delaying recovery from COVID-19). Chart review data was entered into a standardized template capturing all elements reported in results.

Data for specific comorbidities were available for the overall VOMC cohort; these were extracted by data pull for comparison to the comorbidities obtained by chart review for the prolonged symptom cohort. Symptom results were analyzed in Microsoft Excel using descriptive statistics. Comparison between prolonged symptom group and the rest of the VOMC were conducted with Fisher’s exact test using SPSS version 26 (IBM).

## Results

A total of 545 patients were identified as being monitored by VOMC during the study period. 151 were excluded due to symptom onset date not recorded in intake note. 26 patients met study criteria, which constitutes 4.7% of the VOMC cohort.

The majority of patients were female (76.9%), and the median age was 47.5 years [range 23-78]. Racial demographics were as follows: 14 (53.8%) African American, 5 (19.2%) Caucasian, 2 (7.7%) Asian, 5 (19.2%) not recorded. In the overall VOMC cohort, the median age was 48 [range 18-95], females were 67.9% of the population, and African Americans represented 51.0% of the group (Caucasian 20.6%, Asian 3.5%).

Patients with persistent symptoms entered VOMC a median of 9.5 days after symptom onset [range 4-39] and were followed by VOMC for a median of 38 days [range 21-49]. From symptom onset to discharge from VOMC was a median of 47.5 days [range 42-80]. At discharge 24 (92.3%) patients reported significant improvement of symptoms with only 7 (26.9%) reporting that they were back to baseline health (symptom free).

The presence of symptoms in week 5 to 6 are presented in table 1. Respiratory symptoms were most common, reported in 24 patients total (92.3%). The most common reported symptoms were cough, shortness of breath with exertion, sinus congestion and chest tightness. Other common symptoms include fatigue (17 patients, 65%) and headache (13 patients, 50%). Less commonly reported, 9 patients (34.6%) had persistent gastrointestinal symptoms, 6 patients (23%) complained of palpitations, and 3 had persistent low-grade fevers. Of note, 18 (69.2%) reported at least 4 concurrent symptoms.

**Table 1:** Symptoms reported by patients in 6 ^th^ week

|  |  |
| --- | --- |
| **General** | **20 (76.9%)** |
| Fever | 3 (11.5%) |
| Chills | 3 (11.5%) |
| Body aches | 7 (26.9%) |
| Fatigue | 17 (65.4%) |
| Weakness | 7 (26.9%) |
| Joint pain | 8 (30.8) |
| Sweats | 2 (7.7%) |
| **Neurologic** | **20 (76.9%)** |
| Confusion | 1 (3.8%) |
| Dizziness | 3 (11.5%) |
| Headache | 13 (50%) |
| Memory loss | 4 (15.4%) |
| Sleep disturbance | 7 (26.9%) |
| Anxiety | 8 (30.8) |
| **Respiratory** | **24 (92.3%)** |
| Sore throat | 5 (19.2%) |
| Sinus congestion | 12 (46.2%) |
| Loss of smell | 8 (30.8) |
| Altered taste | 1 (3.8%) |
| Cough | 14 (53.8%) |
| Shortness of breath at rest | 3 (11.5%) |
| Shortness of breath with exertion | 13 (50%) |
| Wheezing | 3 (11.5%) |
| Chest tightness | 10 (42.3%) |
| **Gastrointestinal** | **9 (34.6%)** |
| Diarrhea | 3 (11.5%) |
| Abdominal Pain | 1 (3.8%) |
| Nausea | 5 (19.2%) |
| Anorexia | 3 (11.5%) |
| **Other** |  |
| Rash | 1 (3.8%) |
| Palpitations | 6 (23.1%) |
| **Total number of symptoms** |  |
| 1-3 | 7 (26.9%) |
| 4-6 | 13 (50%) |
| 7-9 | 3 (11.5%) |
| 10 or more | 2 (7.7%) |

Due to the presence of persistent symptoms, 16 (61.5%) of patients delayed return to work at least five weeks from symptom onset, 17 (65.3%) delayed return to activity. The most common reasons cited for delayed return to work or activity was fatigue or weakness.

Patient comorbidities are reported in table 2 with comparison to the overall VOMC cohort. Patients had an average of 2.7 comorbid conditions with the most common conditions noted to be BMI >30 (53.8%), asthma (42.3%), allergies (34.6%), and hypertension (34.6%).

**Table 2:** Comorbid conditions

| Table 2: Comorbid conditions |  |  |  |  |
| --- | --- | --- | --- | --- |
|  | Prolonged<br>(n=26) | Non-prolonged<br>(n=470*) | Odds ratio | p value |
| Age >60 | 5 (19.2%) | 108 (23.0%) | 0.80 | 0.812 |
| BMI>30 | 14 (53.8%) | 196 (41.7%) | 1.63 | 0.308 |
| Hypertension | 9 (34.6%) | 156 (33.2%) | 1.07 | 1.000 |
| Coronary artery disease | 1 (3.8%) | 19 (4.0%) | 0.95 | 1.000 |
| Diabetes | 1 (3.8%) | 65 (13.8%) | 0.25 | 0.232 |
| Immunosuppression | 1 (3.8%) | 26 (5.5%) | 0.68 | 1.000 |
| Chronic kidney disease | 1 (3.8%) | 14 (3.0%) | 1.30 | 1.000 |
| Lung Disease | 3 (11.5%) | 11 (2.3%) | 5.44 | 0.032 |
| Asthma | 10 (42.3) | 59 (12.6%) | 4.35 | 0.001 |
| Allergies | 9 (34.6%) | NR |  |  |
| Mood Disorder | 6 (23%) | NR |  |  |
| Migraines | 4 (15.4%) | NR |  |  |
| Autoimmune Disorder | 4 (15.4%) | NR |  |  |
| Previous Tobacco use | 5 (19.2%) | NR |  |  |
| Abbreviations: BMI = Body mass index; NR = Not recorded |  |  |  |  |
| *Patients in prolonged cohort and hospitalized prior to VOMC enrollment not included |  |  |  |  |

The majority of patients (73%) underwent further evaluation at one or more sites during week 3-6 after symptom onset, either in emergency room (n=5, 26.3%), respiratory clinic (n=12, 63%), or telemedicine visit with specialist (n=4, 21%) or primary care (n=4, 21%). Common tests included chest X-ray (n=10), chest CT (n=7), labs (n=8), and echocardiogram (n=4). An alternate non-COVID-19 diagnosis was reached only for 1 patient (exacerbation of heart failure following resolution of COVID). A contributing diagnosis with delayed improvement in COVID-19 was suspected for 13 patients (50%), most commonly allergic rhinitis (n=7, 58.3%), followed by asthma (n=5, 41.7%) and bronchiectasis in 2 patients (16.7%).

## Discussion

Our data demonstrate that a subset of patients followed as outpatients for mild COVID-19 will experience persistent symptoms, here defined as a period greater than 5 weeks, that impacts their ability to return to work and activity. The majority of patients had specific documentation of difficulty with usual activity and/or work due to functional limitation, with only 26.5% demonstrating a return to baseline health. We found, however, that most patients had an improving symptom course in the 6^th^ week of symptoms.

In this cohort, the majority of patients sought further evaluation for their symptoms, with only one patient identified to have a non-COVID-19 alternate diagnosis. Importantly, in patients evaluated for persistent symptoms, a contributing atopic diagnosis or chronic lung disease was often suspected, which lead to specific directed treatments. This possible association is also suggested by comparison of comorbidities between the prolonged symptom group and the overall VOMC cohort (table 2): the incidence of asthma and chronic lung disease (coded at intake visit) were more frequent in the persistent symptom cohort. Anecdotally, we note that many providers report that inhaled bronchodilators and corticosteroids are effective for prolonged COVID-19 symptoms (see video: https://youtu.be/ecPjhdVf41k) and our findings in this report strengthen the case for further research.

While respiratory symptoms (cough, dyspnea on exertion, and other) are most common, a variety of symptoms may present across organ systems including neurologic, cardiac, and gastrointestinal. These manifestations merit further investigation as possible evidence of organ-specific dysfunction caused by “mild” COVID-19 in outpatients.

## Limitations

Our data represent a specific population of patient who enrolled in a telemedicine program at a single center and may not be generalizable to other populations. Additionally, the request for ongoing follow-up calls was not standardized across the cohort so we cannot be certain which symptoms or functional status concerns prompted the continuation of care. It is therefore likely that our data does not capture all patients with persistent symptoms. In a separate phone-call survey project (internal data) 7/158 (4.4%) patients who were called after VOMC discharge (median symptom day 63) reported significant symptoms requiring ongoing medical care by primary care of which only one was identified in this study (met criteria for prolonged VOMC care).

The study period (March-May 2020) overlapped with the spring pollen season, which could explain the occurrence of contributing atopic diagnoses in this study. The peak pollen counts, however, occurred March 20 – April 10 [6], and the dates of discharge of our prolonged symptom cohort from VOMC were 4/24/20-6/8/20 (median: 5/18/20); the median dates for the 6^th^ week of symptoms used for our analysis was 5/4/2020-5/11/2020. Furthermore, the provider documentation used in our analysis attributed the symptoms to COVID-19 as the primary active diagnosis in all care plans except for a single “alternate diagnosis” case.

## Conclusion

For a subset of patients with COVID-19 (4.7% in our cohort), symptom duration is more than 5 weeks and impacts their ability to return to work and activity. The most common persistent symptoms are respiratory in nature. These patients are more likely to have underlying allergic and lung conditions than in the general telemedicine VOMC follow-up cohort. Further research is needed to determine the long-term effects of COVID-19 on patients with persistent symptoms.

## Data Availability

Due to the nature of this research, participants of this study did not agree for their data to be shared publicly, so supporting data is not available.

## CONTRIBUTORS

JO and MC contributed to the concept and design of the study. JO performed the chart review. Both authors were involved in data interpretation revised the manuscript critically for important intellectual content and approved the final version of the manuscript.

## ACKNOWLEDGEMENTS

We would like to thank David Tong, MD MPH for his assistance with data extraction.

